# Hybrid rule-based and on-premises LLM pipeline for extracting CMR and CPET metrics from free-text reports in repaired tetralogy of Fallot

**DOI:** 10.64898/2026.01.21.26344531

**Authors:** Izzet T. Akbasli, Kristen L. Beck, Wilson Liou, Sherry Du, Gabriel Nemer, Orkun Baloglu, Samir Q. Latifi, Bradley S. Marino, Samer Albahra, Animesh Tandon

## Abstract

**Background:** Patients with repaired tetralogy of Fallot (rTOF) require lifelong surveillance with cardiovascular magnetic resonance (CMR) and cardiopulmonary exercise testing (CPET). However, results are frequently stored as unstructured free-text reports, hindering large-scale analysis and research.

**Objectives:** This study aimed to develop and evaluate a privacy-preserving hybrid natural language processing pipeline combining regular expressions (regex) and an on-premises large language model (LLM) to accurately extract key CMR and CPET metrics from legacy free-text reports in patients with rTOF.

**Methods:** We retrospectively analyzed 430 CMR and 262 CPET reports (2005–2023) from patients with rTOF. A two-stage hybrid pipeline was implemented: regex rules were applied first, followed by targeted prompting of an on-premises Llama-3.1-8B-Instruct LLM only when regex failed or returned ambiguous results. Performance was compared against regex-only and LLM-only approaches using coverage, precision, recall, and F1-score.

**Results:** In CMR reports, the hybrid pipeline achieved perfect coverage (1.00) and F1-score (1.00) versus 0.98 coverage and 0.93 F1-score with regex alone, while reducing computational cost by ~75% compared with LLM-only. In CPET reports, the hybrid approach improved F1-score from 0.74 (regex alone) to 0.98, with particularly large gains for semantically complex variables (e.g., peak VO □, test termination reason).

**Conclusions:** A hybrid regex–on-premises LLM pipeline provides near-perfect, efficient, and HIPAA-compliant extraction of clinical metrics from unstructured cardiology reports, offering a scalable solution for retrospective research and quality improvement in congenital heart disease.

## Introduction

Patients with Tetralogy of Fallot (TOF) require life-long surveillance with cardiovascular magnetic resonance (CMR) and cardiopulmonary exercise testing (CPET) to assess ventricular volumes and function, pulmonary regurgitation, and exercise capacity [1–5]. Despite calls for structured reporting [3], CMR and CPET testing results are commonly reported using software that generates free-text or lightly-structured reports, often shaped by multiple local templates that have evolved over decades, introducing variability in both format and terminology[6, 7]. The variability in the structure of reports has created a significant barrier to retrospective or multi-center research and quality improvement initiatives, as extracting basic measurements requires time-consuming manual review, which is error-prone and not scalable.

Rule-based approaches such as regular expressions (regex) can partially automate information retrieval and provide value in radiology and cardiology datasets with stable syntax [8]. However, performance declines when synonyms, modifiers, or slight word□order changes appear (e.g., “LVEF 55%” vs. “Left ventricular ejection fraction was 55%”) or when adjacent entities cause values to be misattributed (e.g., capturing ejection fraction value for right ventricular (RV) instead of left ventricular (LV) metrics). Machine learning named entity recognition (NER) models, built on bidirectional encoder representations from transformers (BERT) or similar transformer backbones, overcome many of these limitations and have attained near–human accuracy in clinical NLP tasks [8–12]. However, the implementation of such systems typically depends on thousands of manually annotated sentences, an effort that demands substantial or impossible resources for centers focused on rare conditions such as TOF.

In contrast, large language models (LLMs) trained on extensive collections of natural language data can readily adapt to new tasks—including named entity recognition (NER) in both English and non-English text [10]. For example, large-scale architectures such as OpenAI’s GPT-4, ChatGPT, and Google’s Gemini have achieved strong NER performance in various medical applications without the need for further fine-tuning [13, 14]. However, because most commercial LLMs are hosted on public cloud platforms, they raise concerns under Health Insurance Portability and Accountability Act (HIPAA) and General Data Protection Regulation (GDPR) regarding the handling of protected health information (PHI) [15, 16]. Therefore, several academic health systems have deployed on-premises or intranet LLMs that process PHI entirely within the institutional firewall [14].

To address the need for accurate yet efficient extraction of numeric CMR data from free-text reports, we implemented a two-stage pipeline in which deterministic regex rules run first and an on-premises LLM is invoked only when regex returns no value or conflicting candidates. This ordering preserves PHI (all inference occurs behind the institutional firewall) and materially reduces token usage and wall-clock time relative to an LLM-only baseline. We hypothesized that this hybrid approach would improve F1 scores over standalone methods. We then assessed the accuracy of data extraction with regex only, LLM only, and regex-LLM hybrid pipelines.

## Materials and Methods

### Study Design

In this retrospective study, an on-premises implementation of the open-source Llama-3.1-8B-Instruct model (8-billion parameter instruction-tuned large language model) was employed to safely extract predefined metrics (detailed below) from free-text CMR and CPET reports through an NER task. The protocol was approved by the Cleveland Clinic Institutional Review Board (IRB #23-910; 26 August 2023). This study was reported in accordance with the STROBE guidelines [17].

### Data

As part of an ongoing study, we identified all patients with repaired TOF (rTOF) who underwent CMR and CPET with 1 year of each other, from March 2005 to September 2023. CMR and CPET reports were retrieved from the institution’s EPIC electronic health record by querying the Clarity database with SAP Crystal Reports. We analyzed all 430 CMR and 262 CPET reports that met the inclusion criteria. Patients were excluded if the interval between their CMR and CPET examinations exceeded one year or if they underwent any surgical intervention between the CMR and CPET, or if the corresponding report was not available in the medical record system.

### Feature extraction and prompting

Thirteen CMR and five CPET variables (listed in Supplementary Table 1) were analyzed. The right-ventricular mass was excluded as it appeared in only four reports. The ground-truth labels were manually reviewed by a pediatrician (ITA) with eight years of experience. Typical string patterns for each metric were first characterized, and bespoke regular-expression functions were written to capture them. LLM prompts were then constructed for zero-shot (task description only), one-shot (single annotated example), and few-shot (multiple examples) extraction scenarios [18]. Regex functions and prompts are provided in the supplementary material. All calls to the model were issued through a custom Python interface that integrated the regex layer with LLM query handling and post-processing (available at github.com/turkalpmd/HeaRTNER). In this hybrid regex–LLM pipeline, we did not implement a fully automated controller that dynamically decides between regex and LLM on a per-report basis. Instead, we empirically identified specific variables for which regex performance was poor (notably LVEF, peak VO□, and test termination reason) and pre-specified that these features would be extracted using LLM output rather than regex in the final dataset.

### Model

An on-premises deployment of the Llama 3.1–8B-Instruct model (fp16 precision) was utilized for this study. The model was obtained from the official Llama repository hosted on Hugging Face (https://huggingface.co/meta-llama/Llama-3.1-8B-Instruct). The checkpoint—originating from Meta’s Llama 3.1 foundation model—was executed without any additional fine-tuning. Inference employed fully deterministic decoding (temperature = 0.0; top-k = 1; top-p = 1.0) with the entire 4096-token context window available for both input and output sequences. All computations were performed behind the institutional firewall, and the model was supplied solely with the specific report section under review, thereby matching the information available to human evaluators.

### Performance Evaluation

Performance metrics were quantified with 95% confidence intervals to ensure precision. Requests were sent to the model via its HTTPS API, the returned values were exported as CSV files, and all performance statistics were computed with scikit-learn in an open-source Python 3.11 environment. We evaluated extraction performance using four complementary metrics and reported 95% confidence intervals for each. Coverage was defined as the proportion of ground-truth rows for which the pipeline returned any (non-missing) value. If the original report did not contain the value to begin with (e.g. no pulmonic regurgitant fraction measured due to valve artifact, or no peak VO2 reported due to early termination), those values were excluded from analysis. Precision was the proportion of correctly extracted values among all extracted values (TP/[TP+FP]). Recall was the proportion of correctly extracted values among all reference values (TP/[TP+FN]). The F1 score was computed as the harmonic mean of precision and recall. Pairwise comparisons of model performance were conducted on overall F1 scores using Wilcoxon signed-rank tests, with p values adjusted for multiple comparisons using the Benjamini–Hochberg false discovery rate (FDR) procedure. Detailed measures of numeric fidelity (e.g., MAE, RMSE, and R^2^) are provided in the Supplementary Material.

## Results

### Population description

430 CMR reports from 183 patients and 262 CPET reports from 124 patients with rTOF were retrospectively analyzed. The median age at the time of CMR was 25.8 years [IQR: 17.4–38.6]; for CPET, it was 22.8 years [IQR: 16.6–34.8]. In both report cohorts, the most frequently observed rTOF subtype was unclassified rTOF (see Table 1).

**Table 1.**
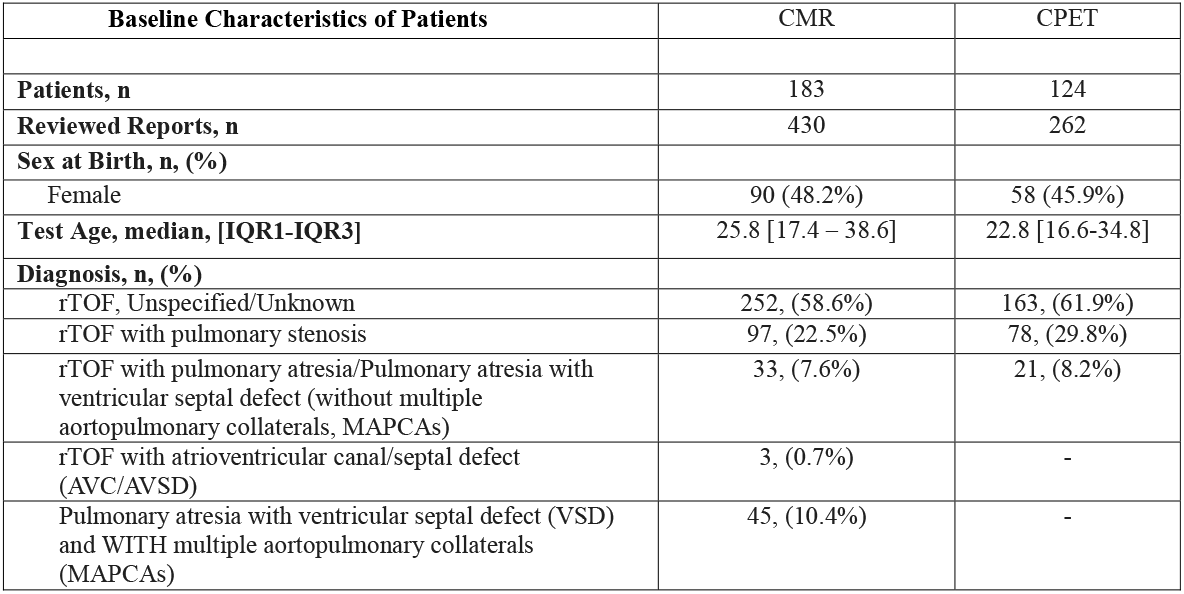
Baseline Characteristics of Patients.

| **Table 1. Baseline Characteristics of Patients** | CMR | CPET |
| --- | --- | --- |
| **Patients, n** | 183 | 124 |
| **Reviewed Reports, n** | 430 | 262 |
| **Sex at Birth, n, (%)** |  |  |
| Female | 90 (48.2%) | 58 (45.9%) |
| **Test Age, median, [IQR1-IQR3]** | 25.8 [17.4 – 38.6] | 22.8 [16.6-34.8] |
| **Diagnosis, n, (%)** |  |  |
| rTOF, Unspecified/Unknown | 252, (58.6%) | 163, (61.9%) |
| rTOF with pulmonary stenosis | 97, (22.5%) | 78, (29.8%) |
| rTOF with pulmonary atresia/Pulmonary atresia with ventricular septal defect (without multiple aortopulmonary collaterals, MAPCAs) | 33, (7.6%) | 21, (8.2%) |
| rTOF with atrioventricular canal/septal defect (AVC/AVSD) | 3, (0.7%) | - |
| Pulmonary atresia with ventricular septal defect (VSD) and WITH multiple aortopulmonary collaterals (MAPCAs) | 45, (10.4%) | - |

Manual chart review revealed a median missing-data rate of 3.9% [IQR: 2.8–5.1] across 13 quantitative CMR variables and 4.2% [IQR: 3.8–4.3] across 5 clinically relevant variables CPET variables (Supplemental Table 1).

### CMR results

In the CMR dataset evaluation, using the LLM, the F1 scores obtained using zero-shot, one-shot, and few-shot prompts (FSP) were 0.87 [95% CI: 0.76–0.94], 0.88 [95% CI: 0.81–0.97], and 0.89 [95% CI: 0.82–0.98], respectively (see Supplementary Table 2). Token usage was observed as 7,644 [95% CI: 7,541–7,803], 10,766 [95% CI: 10,655–10,928], and 13,856 [95% CI: 13,742–13,999] in the same order, with corresponding average response times of 10.0 seconds [95% CI: 9.9–10.0], 7.5 seconds [95% CI: 7.5–7.6], and 7.6 seconds [95% CI: 7.6–7.6]. Considering that the primary objective is to achieve high accuracy, only the results of the FSP models will be presented in subsequent analyses (see Supplementary Table 2).

Fig. 1 illustrates the performance of regex, few□shot LLM, and hybrid pipelines on CMR report extraction, reporting coverage, precision, recall and F1□score with 95% confidence intervals (see Supplementary Table 6). Overall, the regex baseline achieved 0.98 [95% CI: 0.83–0.99] coverage, 0.92 [95% CI: 0.77–0.99] recall, 0.99 [95% CI: 0.94–1.00] precision and an F1□score of 0.93 [95% CI: 0.87–0.99]. Utilizing a standalone few□shot LLM prompt introduced issues in coverage and accuracy: coverage was comparable at 0.97 [95% CI: 0.94–0.99], recall decreased to 0.89 [95% CI: 0.80–0.97], precision decreased to 0.90 [95% CI: 0.85–1.00] and F1□score decreased to 0.89 [95% CI: 0.82–0.98]. In contrast, the hybrid LLM-regex pipeline approached complete extraction, with coverage of 1.00 [95% CI: 1.00–1.00], recall of 1.00 [95% CI: 0.94–1.00], precision of 1.00 [95% CI: 0.94–1.00] and F1□score of 1.00 [95% CI: 0.94–1.00]. Pairwise Wilcoxon signed□rank tests with Benjamini-Hochberg FDR correction confirmed that the hybrid approach significantly outperformed both single□method baselines (regex and FSP) with FDR-adjusted p-values (q) < 0.05 (Supplementary Table 11). Effect sizes were moderate to large, with Cohen’s d ≥ 0.60 and Cliff’s Δ ≥ 0.67 for comparisons involving the hybrid approach.

**Fig. 1.**
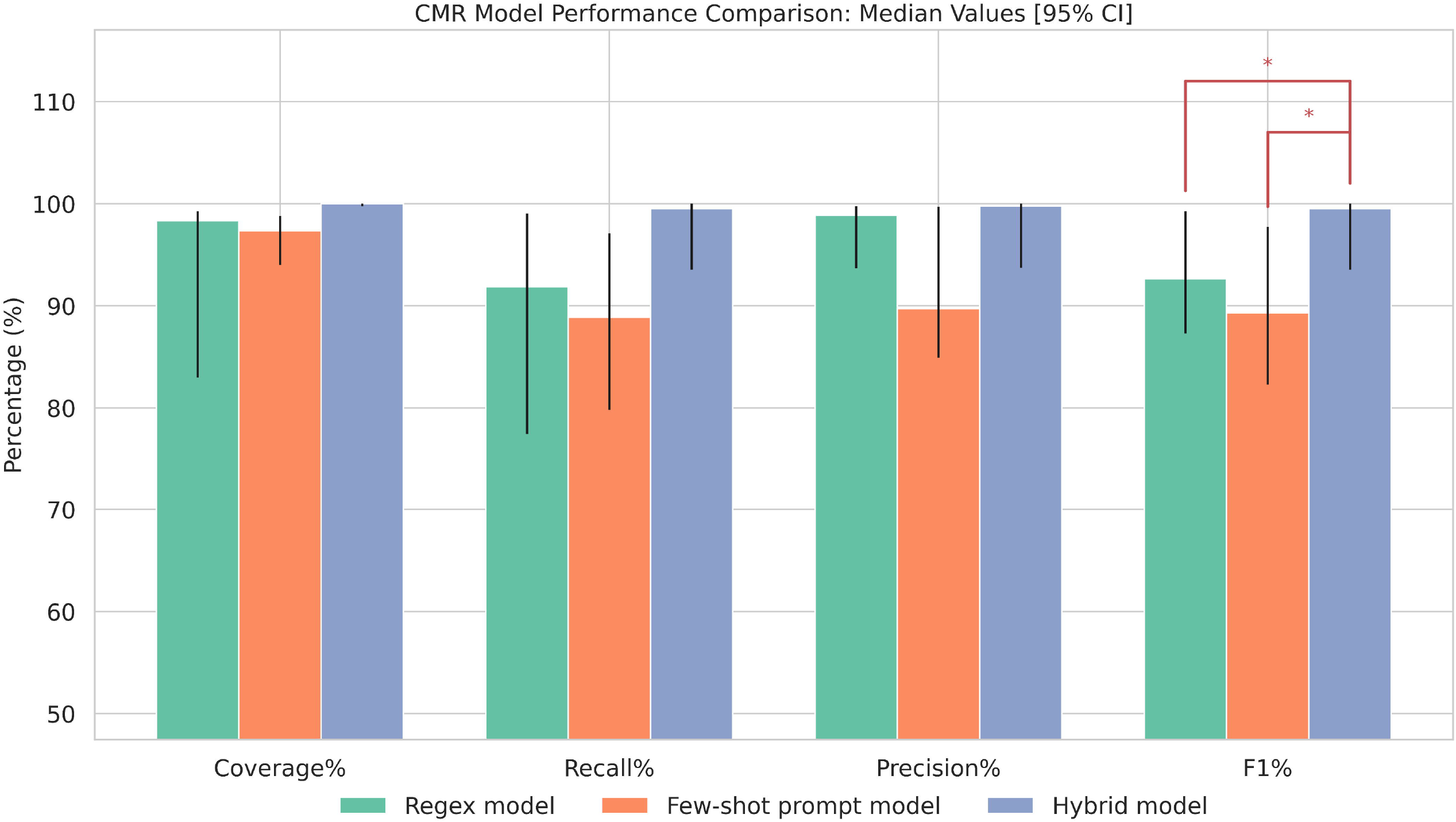
Graph shows performance for the CMR report NER extraction task across three approaches (Regex-only, Few-shot prompt, Hybrid), with median coverage, recall, precision, and F1 (%) ± 95% CI. The hybrid model achieved near perfect coverage and outperformed regex-only and FSP-only methods on all metrics, with statistically significant improvements in F1 score over both alternatives (* indicates p < 0.05).

In terms of specific parameters, although regex achieved reasonably high accuracy for height, weight and body surface area (BSA) with a mean F1-score of 0.88 across these three variables, it retrieved only 23.7% of pulmonic-regurgitant-fraction (PRF) values and misclassified 43% of left-ventricular ejection-fractions (LVEF) as right-ventricular values due to symmetrical lexical patterns (see Supplementary Table 3), as reports often listed ‘ejection fraction’ under both ventricular sections without explicit LVEF/RVEF labels. In contrast, the hybrid workflow increased the median F1 score from regex only of 0.93 up to 1.00 for hybrid while consuming ~75% fewer tokens (≈ 4.5 million) and reducing total runtime by 75.9% (≈ 45 min) relative to the standalone FSP approach (see Supplementary Tables 4 and 5). Detailed variable-level metrics and error audits are provided in the Supplementary Material.

### CPET results

For the desired CPET variables (peak VO□, VE/VCO□ slope, weight, termination reason and test type: treadmill or cycle ergometer), Fig. 2 presents performance of the three extraction pipelines (See Supplementary Table 10). In these tasks, the regex model delivered high coverage 1.00 [95% CI: 1.00–1.00] but relatively low recall 0.74 [95% CI: 0.50–0.88], precision 0.74 [95% CI: 0.11–0.88] and F1□score 0.74 [95% CI: 0.18–0.88]. The few□shot LLM improved these to 1.00 [95% CI: 0.97–1.00], 0.84 [95% CI: 0.70–0.98], 0.88 [95% CI: 0.75–0.98] and 0.86 [95% CI: 0.72–0.98], respectively. The hybrid pipeline further enhanced performance: coverage reached 0.98 [95% CI: 0.95–1.00], recall 0.97 [95% CI: 0.93–0.98], precision 0.98 [95% CI: 0.98–0.99] and F1□score 0.98 [95% CI: 0.96–0.98]. However, pairwise Wilcoxon tests found no statistically significant differences among regex, few□shot and hybrid methods (all p > 0.05), despite large effect sizes in some comparisons (see Supplementary Table 11). Detailed variable-level metrics and error audits are provided in the Supplementary Material.

**Fig. 2.**
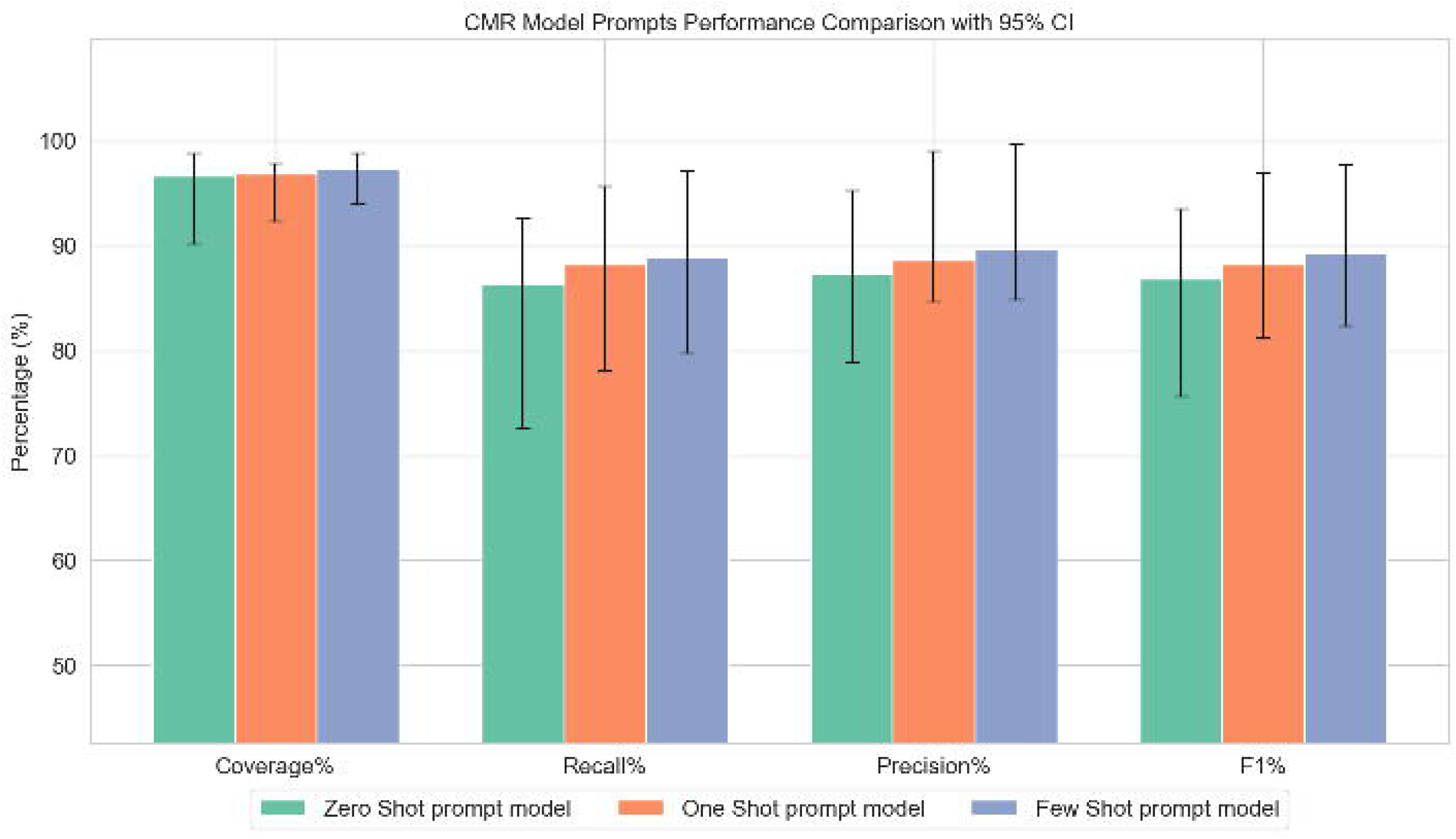
Graph shows performance for the CPET report NER extraction restricted to three variables (peak VO□, VE/VCO□ slope, and weight) across three approaches (regex-only, FSP, hybrid), with mean coverage, recall, precision, and F1 (%) ± 95% CI. The hybrid model achieved near perfect coverage and outperformed regex-only and FSP methods on all metrics.

## Discussion

Overall, this study shows that a two-stage information-extraction strategy, combining rule-based regex with prompting of an on-premises Llama 3.1 LLM, substantially outperforms either method alone when applied to heterogeneous free-text cardiology reports in patients with rTOF. In the CMR reports, the hybrid pipeline increased the median F1 score from 0.93 (regex only) to 1.00 (hybrid) while maintaining near-complete variable coverage (≈98%), and reduced token consumption and end-to-end runtime by roughly 75%. In CPET reports, the hybrid approach notably increased extraction accuracy, raising F1-scores from 0.74 (regex alone) to 0.98, particularly for semantically complex variables such as peak VO□ and test termination reasons.

The CPET results highlighted that where the data are less structured, the LLM and hybrid approaches had significant advantages. Despite coverage being comparable across models, the hybrid approach improved the aggregate F1-score by nearly 30%. Error analysis indicated that the regex system failed on two clinically salient variables. First, inconsistent formatting of peak VO□ values reduced regex performance to an F1 of 0.06 [95% CI 0.04–0.09]. Second, the “test-termination reason” was documented inconsistently as unstructured free text rather than in a standardized field, the regex achieved only an F1 score of 0.18 [95% CI 0.15–0.21] with poor extraction performance and underscoring the necessity of the hybrid approach. The FSP LLM, by contrast, leveraged contextual cues and domain semantics to raise peak VO□ extraction to 0.66 [95% CI 0.61–0.69] and captured termination reason with 0.92 F1 score (95% CI 0.89–0.96]. Incorporating both methods in a hybrid pipeline further improved semantic reasoning, enhancing peak VO□ extraction to 0.97 F1 score while maintaining high accuracy for termination reason. These findings underline the complementary strengths of syntactic precision and semantic comprehension: deterministic rules excel when data are consistently formatted, whereas LLMs interpret context-rich, variably expressed concepts. By uniting both capabilities, the hybrid pipeline offers a practical solution for converting legacy cardiology narratives into high-quality structured datasets, supporting clinical quality-improvement efforts as well as retrospective research.

Structured, database-driven reporting is the ideal solution for consistent data extraction. Although leading cardiology societies have long endorsed standardized nomenclature and reporting templates, the adoption of these frameworks by electronic health record (EHR) systems only commenced around 2010–2015[6, 19]; consequently, many outcome reports produced prior to this period persist as free-text entries in the EHR, reliant on institution-specific templates and clinician dictation[7, 20, 21]. NER can be applied to transform these free-text clinical reports into structured data by identifying and labeling predefined entities. As noted above, when report templates are sufficiently consistent, NER performed through regex can yield high accuracy (F_1_ ≈ 95–97%) at minimal computational cost [22, 23]. However, in less standardized settings, supervised, small□scale language models, such as BERT or ClinicalBERT, become necessary to achieve the same accuracy [10, 11, 22, 23]. These models typically require thousands of annotated examples, a requirement often impractical for rare conditions like TOF.

In terms of data security, our findings demonstrate that high-accuracy clinical NLP can be achieved with a general-purpose, 8-billion-parameter open-source LLM deployed behind the institutional firewall on-premises, without the heavy hardware dependence typically required by larger proprietary models. Because most commercial LLMs are cloud-hosted and thus incompatible with PHI under HIPAA and GDPR unless replicated on costly, security-hardened on-premises clusters [10, 14, 15, 24], we leveraged Cleveland Clinic’s existing infrastructure to deploy the open-source Llama 3.1 model locally. Instead of task-specific fine-tuning, we issued tailored prompts to the pretrained model, avoiding any external transfer of PHI while still achieving strong extraction performance in a limited-annotation setting. Local inference also reduced per-token computational costs compared with cloud services such as OpenAI, Claude, Grok, or Gemini, underscoring the feasibility of privacy-preserving, cost-effective clinical NLP with open-source LLMs in well-resourced healthcare systems [25, 26].

In terms of computational costs, while the hybrid approach in this study prioritizes maximizing data extraction accuracy, applying full LLM inference to every report may not be necessary, or efficient, in future implementations. LLMs, though increasingly reliable, are still prone to hallucinations [13, 27], underscoring the value of rule-based filters to constrain and pre-structure input. Our results suggest that regularly formatted variables can be handled reliably with regex, while more variable or context-dependent fields benefit from LLM processing. Instead of invoking the LLM for every field, future systems could use simple rules to route only the most challenging cases to the model. This selective strategy offers a major advantage: reducing token consumption, runtime, and cost. By reserving LLMs for tasks where they add clear value, and offloading simpler cases to cheaper deterministic tools, future pipelines can become both more scalable and sustainable [26, 28, 29], a priority as automated clinical data extraction becomes more widespread [30, 31].

This study has several limitations. First, it was conducted at a single center and focused on a narrow disease cohort, patients with rTOF, so external validation across multiple institutions and broader patient populations is necessary to establish generalizability. Additionally, reporting styles and documentation practices vary widely between institutions, meaning that the regex patterns developed here may not transfer seamlessly to other sites. As a result, each institution may need to develop its own tailored rule set, which could limit scalability and increase implementation burden. Second, the technical stack used in this study relied on a general-purpose, off-the-shelf LLM without task-specific fine-tuning; while performance was strong, fine-tuned or domain-specific models may offer improved accuracy and reliability especially as semantic complexity increases. Third, the regex filters were not engineered to consistently isolate only the most relevant portions of text before LLM processing, representing a missed opportunity for additional gains in efficiency and cost reduction.

Future research should prioritize the refinement of pre-filtering strategies to dynamically isolate the most relevant text segments for targeted LLM processing. Such approaches could significantly reduce token consumption, enhance computational efficiency, and minimize the likelihood of hallucinations. Concurrently, the development and evaluation of cardiology-specific transformer models fine-tuned for clinical NLP tasks could further improve accuracy, reliability, and safety of automated extraction pipelines [32–36]. Investigating the integration of optimized hybrid NLP workflows directly within EHR systems is another important direction, potentially enabling real-time structured data extraction during patient encounters [31, 37]. This integration would streamline clinical documentation workflows, allowing clinicians to focus on higher-order interpretation and decision-making rather than manual data retrieval [38–41]. Lastly, assessing the environmental impact and computational sustainability of hybrid approaches compared with fully neural, LLM-only methods should be systematically explored, providing evidence-based guidance for institutions aiming to implement scalable, cost-effective, and environmentally responsible clinical NLP solutions [26, 29].

## Conclusion

This study demonstrates that a strategically designed hybrid approach, combining deterministic rule-based extraction and on-premises transformer-based language modeling, delivers high-accuracy data retrieval from heterogeneous free-text cardiology reports, while substantially reducing computational overhead. This method represents an impactful advance toward automating clinical information curation, providing clinicians and researchers alike with reliable, structured data previously locked within decades of legacy reports. Nevertheless, ongoing refinement, including tailored model optimization, robust infrastructure development, and diligent oversight, remains essential to translating these advances into routine clinical practice and collaborative, large-scale research initiatives.

## Supporting information

Supplemental Digital Content

## Acknowledgements

None.

## Statements & Declarations

### Author Contributions

I.T.A. led the conceptualization and methodology, performed formal analysis, wrote the original draft, produced the visualization, and co-administered the project; I.T.A., W.L. and S.A. developed the software; A.T. carried out validation, investigation, resource acquisition, supervision, writing – review & editing, and co-administration; S.D. and G.N. curated the data; W.L. and K.L.B. co-wrote the original draft; and S.Q.L., B.S.M., S.A., K.L.B, and O.B. contributed to writing – review & editing.

### Data availability

All data produced in the present study are available upon reasonable request to the authors.

### Code availability

The custom code used in this study is available at https://github.com/turkalpmd/HeaRTNER

### Ethics approval

The protocol was approved by the Cleveland Clinic Institutional Review Board (IRB #23-910; 26 August 2023)

### Consent to participate

Requirement for informed consent was waived due to the study’s retrospective nature and minimal associated risk to patients.

### Consent to publish

Not applicable

