## Supplemental Digital Content for "Hybrid rule-based and on-premises LLM pipeline for extracting CMR and CPET metrics from free-text reports in repaired tetralogy of Fallot"

**Akbasli et. al.,**

**2025**

[Supplementary Table 6 Summary of tables: Aggregated Median [IQR] and Mean ± SD of Performance Metrics for CMR Models	14](#_Toc214016033)

[Supplementary Table 10 Summary of tables: Aggregated Median [IQR] and Mean ± SD of Performance Metrics for CPET Models	18](#_Toc214016038)

### STROBE Statement—checklist of items that should be included in reports of observational studies

|  | Item No. | Recommendation | Page  No. |
| --- | --- | --- | --- |
| **Title and abstract** | 1 | (*a*) Indicate the study’s design with a commonly used term in the title or the abstract | 1 |
|  |  | (*b*) Provide in the abstract an informative and balanced summary of what was done and what was found | 1 |
| Introduction | | | |
| Background/rationale | 2 | Explain the scientific background and rationale for the investigation being reported | 4 |
| Objectives | 3 | State specific objectives, including any prespecified hypotheses | 4-5 |
| Methods | | | |
| Study design | 4 | Present key elements of study design early in the paper | 5 |
| Setting | 5 | Describe the setting, locations, and relevant dates, including periods of recruitment, exposure, follow-up, and data collection | 5-6 |
| Participants | 6 | (*a*) *Cohort study*—Give the eligibility criteria, and the sources and methods of selection of participants. Describe methods of follow-up  *Case-control study*—Give the eligibility criteria, and the sources and methods of case ascertainment and control selection. Give the rationale for the choice of cases and controls  *Cross-sectional study*—Give the eligibility criteria, and the sources and methods of selection of participants | 5-6 |
|  |  | (*b*) *Cohort study*—For matched studies, give matching criteria and number of exposed and unexposed  *Case-control study*—For matched studies, give matching criteria and the number of controls per case | N/A |
| Variables | 7 | Clearly define all outcomes, exposures, predictors, potential confounders, and effect modifiers. Give diagnostic criteria, if applicable | 6 |
| Data sources/ measurement | 8* | For each variable of interest, give sources of data and details of methods of assessment (measurement). Describe comparability of assessment methods if there is more than one group | *6* |
| Bias | 9 | Describe any efforts to address potential sources of bias | 6-7 |
| Study size | 10 | Explain how the study size was arrived at | N/A |
| Quantitative variables | 11 | Explain how quantitative variables were handled in the analyses. If applicable, describe which groupings were chosen and why | 6-7 |
| Statistical methods | 12 | (*a*) Describe all statistical methods, including those used to control for confounding | 7 |
|  |  | (*b*) Describe any methods used to examine subgroups and interactions | 7 |
|  |  | (*c*) Explain how missing data were addressed | 6-7 |
|  |  | (*d*) *Cohort study*—If applicable, explain how loss to follow-up was addressed  *Case-control study*—If applicable, explain how matching of cases and controls was addressed  *Cross-sectional study*—If applicable, describe analytical methods taking account of sampling strategy | N/A |
|  |  | (*e*) Describe any sensitivity analyses |  |
| Results |  |  |  |
| Participants | 13* | (a) Report numbers of individuals at each stage of study—eg numbers potentially eligible, examined for eligibility, confirmed eligible, included in the study, completing follow-up, and analysed | 7 |
|  |  | (b) Give reasons for non-participation at each stage | 7 |
|  |  | (c) Consider use of a flow diagram | N/A |
| Descriptive data | 14* | (a) Give characteristics of study participants (eg demographic, clinical, social) and information on exposures and potential confounders | 8-9 |
|  |  | (b) Indicate number of participants with missing data for each variable of interest | N/A |
|  |  | (c) *Cohort study*—Summarise follow-up time (eg, average and total amount) | N/A |
| Outcome data | 15* | *Cohort study*—Report numbers of outcome events or summary measures over time | N/A |
|  |  | *Case-control study—*Report numbers in each exposure category, or summary measures of exposure | N/A |
|  |  | *Cross-sectional study—*Report numbers of outcome events or summary measures | N/A |
| Main results | 16 | (*a*) Give unadjusted estimates and, if applicable, confounder-adjusted estimates and their precision (eg, 95% confidence interval). Make clear which confounders were adjusted for and why they were included | 7-11 |
|  |  | (*b*) Report category boundaries when continuous variables were categorized | N/A |
|  |  | (*c*) If relevant, consider translating estimates of relative risk into absolute risk for a meaningful time period | N/A |
| Other analyses | 17 | Report other analyses done—eg analyses of subgroups and interactions, and sensitivity analyses | 7,10-11 |
| Discussion |  |  |  |
| Key results | 18 | Summarise key results with reference to study objectives | 11-12 |
| Limitations | 19 | Discuss limitations of the study, taking into account sources of potential bias or imprecision. Discuss both direction and magnitude of any potential bias | 12-13 |
| Interpretation | 20 | Give a cautious overall interpretation of results considering objectives, limitations, multiplicity of analyses, results from similar studies, and other relevant evidence | 14 |
| Generalisability | 21 | Discuss the generalisability (external validity) of the study results | 14 |
| Funding | 22 | Give the source of funding and the role of the funders for the present study and, if applicable, for the original study on which the present article is based | 15 |

*Give information separately for cases and controls in case-control studies and, if applicable, for exposed and unexposed groups in cohort and cross-sectional studies.

**Note:** An Explanation and Elaboration article discusses each checklist item and gives methodological background and published examples of transparent reporting. The STROBE checklist is best used in conjunction with this article (freely available on the Web sites of PLoS Medicine at http://www.plosmedicine.org/, Annals of Internal Medicine at http://www.annals.org/, and Epidemiology at http://www.epidem.com/). Information on the STROBE Initiative is available at www.strobe-statement.org.

### Description of the features

#### Supplementary Table 1 - Descriptive Statistics and Missingness of CMR and CPET Variables

| **Candidate Feature** | **Available Rows** | **Missing Ratio_%** | **Min** | **Max** | **Mean** | **Std** | **Median** | **IQR Q1** | **IQR Q3** |
| --- | --- | --- | --- | --- | --- | --- | --- | --- | --- |
| **CMR Features**  **(n = 430)** |  |  |  |  |  |  |  |  |  |
| **Height** | 408 | 5.12 | 0.9 | 193 | 162.26 | 21.28 | 165 | 157.5 | 173 |
| **Weight** | 408 | 5.12 | 0.9 | 174.6 | 73.97 | 25.88 | 70.8 | 58.08 | 90.7 |
| **BSA** | 423 | 1.63 | 0.59 | 71 | 1.95 | 3.39 | 1.78 | 1.58 | 2.02 |
| **LVEDV** | 412 | 4.19 | 45 | 1166 | 154.68 | 73.14 | 144 | 119 | 177.25 |
| **LVESV** | 413 | 3.95 | 16 | 277 | 70.55 | 35.91 | 62 | 49 | 81 |
| **LVMassED** | 327 | 23.95 | 24 | 281 | 95.86 | 33.01 | 91 | 73 | 113 |
| **LVStrokeVolume** | 413 | 3.95 | 29 | 1113 | 83.71 | 55.8 | 79 | 66 | 95 |
| **LVEF** | 416 | 3.26 | 21 | 95 | 54.95 | 7.76 | 56 | 51 | 60 |
| **RVEDV** | 418 | 2.79 | 76 | 516 | 216.95 | 66.37 | 213 | 172 | 248 |
| **RVESV** | 418 | 2.79 | 26 | 319 | 116.73 | 45.66 | 112 | 86.25 | 136.5 |
| **RVStrokeVolume** | 416 | 3.26 | 26 | 229 | 100.07 | 28.95 | 96 | 79 | 116.25 |
| **RVEF** | 422 | 1.86 | 14 | 77 | 46.85 | 7.61 | 47 | 42 | 52 |
| **PRF** | 396 | 7.91 | 0 | 77 | 26.86 | 19.44 | 28 | 8 | 43 |
| **CPET Features**  **(n = 263)** |  |  |  |  |  |  |  |  |  |
| **peakVO₂** | 249 | 4.96 | 8.5 | 53.2 | 26.69 | 7.77 | 25.9 | 21.7 | 31.2 |
| **VE/VCO2slope** | 229 | 12.6 | 17.4 | 47.9 | 28.95 | 4.54 | 28.5 | 26 | 31 |
| **Weight** | 261 | 0.38 | 23 | 172.32 | 72.35 | 24.04 | 68.1 | 55.8 | 88 |
| **TEST TYPE*** | 250 | 4.94 | NA | NA | NA | NA | NA | NA | NA |

**Test type* was the only categorical CPET variable; among available reports, treadmill tests accounted for **96%** (cycle ergometer **4%**). Abbreviations: BSA, body surface area; LVEDV, left-ventricular end-diastolic volume; LVESV, left-ventricular end-systolic volume; LVSV, left-ventricular stroke volume; LVEF, left-ventricular ejection fraction; LVMassED, left-ventricular mass at end-diastole; RVEDV, right-ventricular end-diastolic volume; RVESV, right-ventricular end-systolic volume; RVSV, right-ventricular stroke volume; RVEF, right-ventricular ejection fraction; PRF, pulmonic regurgitant fraction; peak VO₂, peak oxygen consumption; VE/VCO₂ slope, minute-ventilation to carbon-dioxide production slope.

### Comparison of the prompts models

#### Supplementary Table 2 Prompt Comparisons for CMR Reports

|  | ZERO SHOT PROMPT |  | ONE SHOT PROMPT | | few SHOT PROMPT | |
| --- | --- | --- | --- | --- | --- | --- |
|  | Median  [IQR1-IQR3] | Mean  ± SD | Median  [IQR1-IQR3] | Mean  ± SD | Median  [IQR1-IQR3] | Mean  ± SD |
| Coverage (%) | 96.61  [90.14 – 98.8] | 91.69  ± 10.89 | 96.84  [92.31 – 97.85] | 91.44  ± 10.94 | 97.33  [93.99 – 98.8] | 91.97  ± 11.02 |
| Correct (%) | 86.3  [72.6 – 92.58] | 80.08  ± 17.8 | 88.22  [78.12 – 95.69] | 81.96  ± 17.55 | 88.86  [79.81 – 97.09] | 83.29  ± 18.05 |
| Recall (%) | 86.3  [72.6 – 92.58] | 80.08  ± 17.8 | 88.22  [78.12 – 95.69] | 81.96  ± 17.55 | 88.86  [79.81 – 97.09] | 83.29  ± 18.05 |
| Precision (%) | 87.35  [78.9 – 95.29] | 86.35  ± 10.82 | 88.57  [84.64 – 99.0] | 88.66  ± 10.57 | 89.71  [84.91 – 99.69] | 89.55  ± 11.08 |
| F1 (%) | 86.82  [75.69 – 93.54] | 82.84  ± 14.79 | 88.22  [81.25 – 96.95] | 84.92  ± 14.6 | 89.29  [82.28 – 97.71] | 86.04  ± 15.08 |
| Token Count | 7644  [7541–7803] | 8305  ± 1905 | 10766  [10655-10928] | 11427  ± 1905 | 13856  [13742-1399] | 14517  ± 1905 |
| Time (second) | 10  [9.95-10.07] | 7.66  ±0.37 | 7.55  [7.51-7.60] | 10.2  ±0.74 | 7.62  [7.59-7.68] | 7.77  ± 0.43 |

#### Supplementary Figure 1

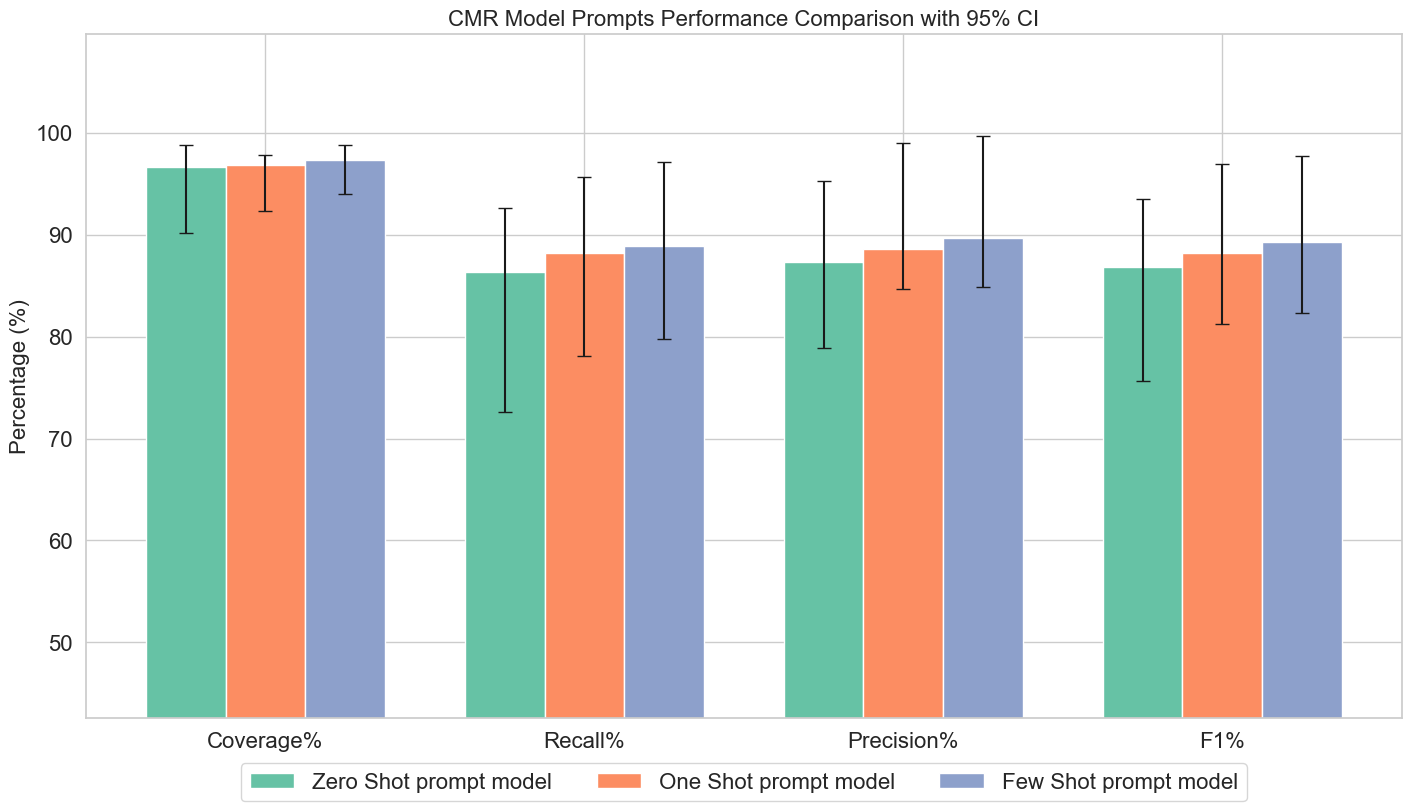

**Supplementary Figure 1:** Graph shows performance for the CMR report NER extraction task across three prompt approaches (zero shot prompt, one shot prompt, few shot prompt), with median Coverage, Recall, Precision, and F₁ (%) ± 95% CI. The hybrid model achieved near perfect coverage and outperformed RegEx-only and FSP methods on all metrics.

### CMR Models Performance: Performance Metrics for Individual CMR Features Across Three Model Configurations

#### Supplementary Table 3 RegEx Model Performance Metrics

| Regex MODEL | Coverage (%) | Correct (%) | Recall (%) | Precision (%) | F1 (%) |
| --- | --- | --- | --- | --- | --- |
| Height | 77.21 [73.14, 81.28] | 77.21 [73.14, 81.28] | 77.21 [73.14, 81.28] | 100.0 [100.0, 100.0] | 87.14 [84.12, 89.37] |
| Weight | 77.45 [73.4, 81.51] | 77.45 [73.4, 81.51] | 77.45 [73.4, 81.51] | 100.0 [100.0, 100.0] | 87.29 [84.28, 89.58] |
| BSA | 82.98 [79.4, 86.56] | 82.03 [78.37, 85.69] | 82.03 [78.37, 85.69] | 98.86 [97.75, 99.97] | 89.66 [87.02, 91.43] |
| LVEDV | 99.51 [98.84, 100.0] | 99.03 [98.08, 99.98] | 99.03 [98.08, 99.98] | 99.51 [98.84, 100.0] | 99.27 [98.18, 99.55] |
| LVESV | 99.27 [98.45, 100.0] | 99.03 [98.09, 99.98] | 99.03 [98.09, 99.98] | 99.76 [99.28, 100.0] | 99.39 [98.42, 99.7] |
| LVMassED | 100.0 [100.0, 100.0] | 100.0 [100.0, 100.0] | 100.0 [100.0, 100.0] | 100.0 [100.0, 100.0] | 100.0 [99.11, 99.97] |
| LVStrokeVolume | 99.27 [98.45, 100.0] | 99.03 [98.09, 99.98] | 99.03 [98.09, 99.98] | 99.76 [99.28, 100.0] | 99.39 [98.42, 99.66] |
| LVEF | 93.03 [90.58, 95.48] | 50.96 [46.16, 55.77] | 50.96 [46.16, 55.77] | 54.78 [49.82, 59.74] | 52.8 [49.2, 56.26] |
| RVEDV | 98.33 [97.1, 99.56] | 91.87 [89.25, 94.49] | 91.87 [89.25, 94.49] | 93.43 [91.04, 95.83] | 92.64 [90.32, 94.1] |
| RVESV | 98.33 [97.1, 99.56] | 92.11 [89.52, 94.69] | 92.11 [89.52, 94.69] | 93.67 [91.32, 96.03] | 92.88 [90.82, 94.33] |
| RVStrokeVolume | 98.8 [97.75, 99.85] | 92.55 [90.02, 95.07] | 92.55 [90.02, 95.07] | 93.67 [91.32, 96.03] | 93.11 [91.14, 94.57] |
| RVEF | 92.18 [89.62, 94.74] | 86.73 [83.49, 89.97] | 86.73 [83.49, 89.97] | 94.09 [91.74, 96.43] | 90.26 [87.84, 92.11] |
| PRF | 23.74 [19.55, 27.93] | 14.65 [11.16, 18.13] | 14.65 [11.16, 18.13] | 61.7 [51.87, 71.53] | 23.67 [19.63, 28.39] |
| Median [IQR] | 98.33 [82.98 – 99.27] | 91.87 [77.45 – 99.03] | 91.87 [77.45 – 99.03] | 98.86 [93.67 – 99.76] | 92.64 [87.29 – 99.27] |
| Mean ± SD | 87.7 ± 20.98 | 81.74 ± 24.27 | 81.74 ± 24.27 | 91.48 ± 15.08 | 85.19 ± 22.15 |

For each CMR feature—height, weight, body surface area (BSA), left ventricular end diastolic volume (LVEDV), left ventricular end systolic volume (LVESV), left ventricular mass (LVMASSED), left ventricular stroke volume (LVSV), left ventricular ejection fraction (LVEF), right ventricular end diastolic volume (RVEDV), right ventricular end systolic volume (RVESV), right ventricular stroke volume (RVSV), right ventricular ejection fraction (RVEF), and pulmonary regurgitation fraction (PRF)—performance metrics (coverage, correct, recall, precision, and F₁) are presented separately for the regex model, few-shot model, and hybrid model. Values are reported as median [interquartile range] and mean ± standard deviation. FSP models means Few-shot prompt model.

| FSP Model | Coverage (%) | Correct (%) | Recall (%) | Precision (%) | F1 (%) |
| --- | --- | --- | --- | --- | --- |
| Height | 70.1 [65.66, 74.54] | 52.45 [47.61, 57.3] | 52.45 [47.61, 57.3] | 74.83 [69.8, 79.86] | 61.67 [57.76, 65.25] |
| Weight | 69.12 [64.63, 73.6] | 49.75 [44.9, 54.61] | 49.75 [44.9, 54.61] | 71.99 [66.74, 77.23] | 58.84 [55.13, 62.63] |
| BSA | 81.32 [77.61, 85.04] | 57.92 [53.21, 62.62] | 57.92 [53.21, 62.62] | 71.22 [66.44, 76.01] | 63.89 [60.59, 67.02] |
| LVEDV | 97.33 [95.77, 98.89] | 97.09 [95.46, 98.71] | 97.09 [95.46, 98.71] | 99.75 [99.26, 100.0] | 98.4 [97.14, 99.0] |
| LVESV | 95.4 [93.38, 97.42] | 95.16 [93.09, 97.23] | 95.16 [93.09, 97.23] | 99.75 [99.25, 100.0] | 97.4 [95.74, 98.18] |
| LVMassED | 100.0 [100.0, 100.0] | 99.69 [99.1, 100.0] | 99.69 [99.1, 100.0] | 99.69 [99.1, 100.0] | 99.69 [98.64, 99.84] |
| LVStrokeVolume | 97.34 [95.78, 98.89] | 84.75 [81.28, 88.21] | 84.75 [81.28, 88.21] | 87.06 [83.78, 90.35] | 85.89 [83.21, 88.08] |
| LVEF | 100.0 [100.0, 100.0] | 88.22 [85.12, 91.32] | 88.22 [85.12, 91.32] | 88.22 [85.12, 91.32] | 88.22 [85.83, 89.99] |
| RVEDV | 98.8 [97.76, 99.85] | 97.13 [95.53, 98.73] | 97.13 [95.53, 98.73] | 98.31 [97.06, 99.55] | 97.71 [96.32, 98.37] |
| RVESV | 95.45 [93.46, 97.45] | 94.26 [92.03, 96.49] | 94.26 [92.03, 96.49] | 98.75 [97.66, 99.84] | 96.45 [94.79, 97.41] |
| RVStrokeVolume | 93.99 [91.71, 96.27] | 79.81 [75.95, 83.67] | 79.81 [75.95, 83.67] | 84.91 [81.36, 88.46] | 82.28 [79.35, 84.64] |
| RVEF | 99.05 [98.13, 99.98] | 88.86 [85.86, 91.86] | 88.86 [85.86, 91.86] | 89.71 [86.8, 92.63] | 89.29 [86.85, 91.08] |
| PRF | 97.73 [96.26, 99.2] | 97.73 [96.26, 99.2] | 97.73 [96.26, 99.2] | 100.0 [100.0, 100.0] | 98.85 [97.71, 99.3] |
| Median [IQR] | 97.33 [93.99 – 98.8] | 88.86 [79.81 – 97.09] | 88.86 [79.81 – 97.09] | 89.71 [84.91 – 99.69] | 89.29 [82.28 – 97.71] |
| Mean ± SD | 91.97 ± 11.02 | 83.29 ± 18.05 | 83.29 ± 18.05 | 89.55 ± 11.08 | 86.04 ± 15.08 |

#### Supplementary Table 4 Few Shot Prompts (FSP) Model Performance Metrics

For each CMR feature—height, weight, body surface area (BSA), left ventricular end diastolic volume (LVEDV), left ventricular end systolic volume (LVESV), left ventricular mass (LVMASSED), left ventricular stroke volume (LVSV), left ventricular ejection fraction (LVEF), right ventricular end diastolic volume (RVEDV), right ventricular end systolic volume (RVESV), right ventricular stroke volume (RVSV), right ventricular ejection fraction (RVEF), and pulmonary regurgitation fraction (PRF)—performance metrics (coverage, correct, recall, precision, and F₁) are presented separately for the regex model, few-shot model, and hybrid model. Values are reported as median [interquartile range] and mean ± standard deviation. FSP models means Few-shot prompt model.

| Hybrid Model | Coverage (%) | Correct (%) | Recall (%) | Precision (%) | F1 (%) |
| --- | --- | --- | --- | --- | --- |
| Height | 100.0 [100.0, 100.0] | 100.0 [100.0, 100.0] | 100.0 [100.0, 100.0] | 100.0 [100.0, 100.0] | 100.0 [99.3, 99.97] |
| Weight | 100.0 [100.0, 100.0] | 100.0 [100.0, 100.0] | 100.0 [100.0, 100.0] | 100.0 [100.0, 100.0] | 100.0 [99.35, 99.98] |
| BSA | 100.0 [100.0, 100.0] | 100.0 [100.0, 100.0] | 100.0 [100.0, 100.0] | 100.0 [100.0, 100.0] | 100.0 [99.36, 99.98] |
| LVEDV | 100.0 [100.0, 100.0] | 99.51 [98.84, 100.0] | 99.51 [98.84, 100.0] | 99.51 [98.84, 100.0] | 99.51 [98.61, 99.7] |
| LVESV | 99.52 [98.85, 100.0] | 99.27 [98.45, 100.0] | 99.27 [98.45, 100.0] | 99.76 [99.28, 100.0] | 99.51 [98.59, 99.73] |
| LVMassED | 100.0 [100.0, 100.0] | 100.0 [100.0, 100.0] | 100.0 [100.0, 100.0] | 100.0 [100.0, 100.0] | 100.0 [99.22, 99.96] |
| LVStrokeVolume | 99.76 [99.28, 100.0] | 99.52 [98.85, 100.0] | 99.52 [98.85, 100.0] | 99.76 [99.28, 100.0] | 99.64 [98.73, 99.81] |
| LVEF | 100.0 [100.0, 100.0] | 99.52 [98.85, 100.0] | 99.52 [98.85, 100.0] | 99.52 [98.85, 100.0] | 99.52 [98.58, 99.74] |
| RVEDV | 100.0 [100.0, 100.0] | 93.06 [90.63, 95.5] | 93.06 [90.63, 95.5] | 93.06 [90.63, 95.5] | 93.06 [90.93, 94.49] |
| RVESV | 100.0 [100.0, 100.0] | 93.54 [91.18, 95.9] | 93.54 [91.18, 95.9] | 93.54 [91.18, 95.9] | 93.54 [91.5, 94.93] |
| RVStrokeVolume | 99.52 [98.85, 100.0] | 93.27 [90.86, 95.68] | 93.27 [90.86, 95.68] | 93.72 [91.38, 96.06] | 93.49 [91.68, 94.78] |
| RVEF | 100.0 [100.0, 100.0] | 100.0 [100.0, 100.0] | 100.0 [100.0, 100.0] | 100.0 [100.0, 100.0] | 100.0 [99.35, 99.97] |
| PRF | 97.98 [96.59, 99.37] | 88.89 [85.79, 91.98] | 88.89 [85.79, 91.98] | 90.72 [87.83, 93.61] | 89.8 [87.53, 91.51] |
| Median [IQR] | 100.0 [99.76 – 100.0] | 99.52 [93.54 – 100.0] | 99.52 [93.54 – 100.0] | 99.76 [93.72 – 100.0] | 99.52 [93.54 – 100.0] |
| Mean ± SD | 99.75 ± 0.56 | 97.43 ± 3.81 | 97.43 ± 3.81 | 97.66 ± 3.47 | 97.54 ± 3.64 |

#### Supplementary Table 5 Hybrid Model Performance Metrics

For each CMR feature—height, weight, body surface area (BSA), left ventricular end diastolic volume (LVEDV), left ventricular end systolic volume (LVESV), left ventricular mass (LVMASSED), left ventricular stroke volume (LVSV), left ventricular ejection fraction (LVEF), right ventricular end diastolic volume (RVEDV), right ventricular end systolic volume (RVESV), right ventricular stroke volume (RVSV), right ventricular ejection fraction (RVEF), and pulmonary regurgitation fraction (PRF)—performance metrics (coverage, correct, recall, precision, and F₁) are presented separately for the regex model, few-shot model, and hybrid model. Values are reported as median [interquartile range] and mean ± standard deviation. FSP models means Few-shot prompt model.

#### Supplementary Table 6 Summary of tables: Aggregated Median [IQR] and Mean ± SD of Performance Metrics for CMR Models

|  | Regex Model | | FSP Model | | hybrid model | |
| --- | --- | --- | --- | --- | --- | --- |
|  | Median  [IQR1-IQR3] | Mean  ± SD | Median  [IQR1-IQR3] | Mean  ± SD | Median  [IQR1-IQR3] | Mean  ± SD |
| Coverage (%) | 98.33  [82.98 – 99.27] | 87.7  ± 20.98 | 97.33  [93.99 – 98.8] | 91.97  ± 11.02 | 100.0  [99.76 – 100.0] | 99.75  ± 0.56 |
| Correct (%) | 91.87  [77.45 – 99.03] | 81.74  ± 24.27 | 88.86  [79.81 – 97.09] | 83.29  ± 18.05 | 99.52  [93.54 – 100.0] | 97.43  ± 3.81 |
| Recall (%) | 91.87  [77.45 – 99.03] | 81.74  ± 24.27 | 88.86  [79.81 – 97.09] | 83.29  ± 18.05 | 99.52  [93.54 – 100.0] | 97.43  ± 3.81 |
| Precision (%) | 98.86  [93.67 – 99.76] | 91.48  ± 15.08 | 89.71  [84.91 – 99.69] | 89.55  ± 11.08 | 99.76  [93.72 – 100.0] | 97.66  ± 3.47 |
| F1 (%) | 92.64  [87.29 – 99.27] | 85.19  ± 22.15 | 89.29  [82.28 – 97.71] | 86.04  ± 15.08 | 99.52  [93.54 – 100.0] | 97.54  ± 3.64 |

Coverage, Correct, Recall, Precision, and F₁ scores are reported as median [interquartile range (IQR₁–IQR₃)] and mean ± standard deviation (SD) for each model configuration: RegEx, few-shot, and hybrid. Abbreviation: CMR = cardiac magnetic resonance, FSP = Few-shot prompt model.

### CPET Models Performance: Performance Metrics for Individual CPET Features Across Three Model Configurations

| Regex Model | Coverage (%) | Correct (%) | Recall (%) | Precision (%) | F1 (%) |
| --- | --- | --- | --- | --- | --- |
| peakVO₂ | 85.94 [81.63, 90.26] | 6.02 [3.07, 8.98] | 6.02 [3.07, 8.98] | 7.01 [3.59, 10.43] | 6.48 [4.56, 9.02] |
| VE/VCO2slope | 100.0 [100.0, 100.0] | 88.21 [84.03, 92.39] | 88.21 [84.03, 92.39] | 88.21 [84.03, 92.39] | 88.21 [85.0, 90.76] |
| Weight | 100.0 [100.0, 100.0] | 98.47 [96.98, 99.96] | 98.47 [96.98, 99.96] | 98.47 [96.98, 99.96] | 98.47 [96.68, 99.13] |
| TEST TYPE | 100.0 [100.0, 100.0] | 74.43 [69.14, 79.71] | 74.43 [69.14, 79.71] | 74.43 [69.14, 79.71] | 74.43 [70.82, 77.79] |
| TERMINATION REASON | 100.0 [96.8, 100.0] | 47.3 [43.9, 50.6] | 50.0 [44.1, 55.7] | 11.6 [9.7, 13.8] | 18.0 [15.2, 21.7] |
| Median [IQR] | 100.0 [100.0, 100.0] | 74.43 [47.33, 88.21] | 74.43 [50.0, 88.21] | 74.43 [11.68, 88.21] | 74.43 [18.00, 88.21] |
| Mean ± SD | 97.19 ± 6.29 | 62.89 ± 37.14 | 63.43 ± 36.88 | 55.94 ± 43.45 | 57.12 ± 42.04 |

#### Supplementary Table 7 RegEx Model Performance Metrics

For each CPET feature—peak V̇O₂, VE/VCO₂ slope, and weight—performance metrics (coverage, correct, recall, precision, and F₁) are reported separately for the regex model, few-shot prompting (FSP) model, and hybrid model. Values are presented as median [interquartile range] and mean ± standard deviation. FSP models means Few-shot prompt model.

#### Supplementary Table 8 FSP Model Performance Metrics

| FSP Model | Coverage (%) | Correct (%) | Recall (%) | Precision (%) | F1 (%) |
| --- | --- | --- | --- | --- | --- |
| peakVO₂ | 99.6 [98.81, 100.0] | 65.46 [59.56, 71.37] | 65.46 [59.56, 71.37] | 65.73 [59.82, 71.63] | 65.59 [61.3, 69.44] |
| VE/VCO2slope | 90.83 [87.09, 94.57] | 71.18 [65.31, 77.05] | 71.18 [65.31, 77.05] | 78.37 [72.77, 83.96] | 74.6 [70.2, 78.45] |
| Weight | 100.0 [100.0, 100.0] | 98.47 [96.98, 99.96] | 98.47 [96.98, 99.96] | 98.47 [96.98, 99.96] | 98.47 [96.74, 99.08] |
| Test TYPE | 100.0 [100.0, 100.0] | 97.71 [95.9, 99.52] | 97.71 [95.9, 99.52] | 97.71 [95.9, 99.52] | 97.71 [95.92, 98.62] |
| Termination reason | 97.8 [95.4 – 99.2] | 89.3 [85.0 – 93.6] | 90.1 [87.0 – 93.2] | 94.8 [92.1 – 97.5] | 92.2 [89.6 – 96.3] |
| Median [IQR]* | 100.0 [96.48, 100.0] | 81.32 [57.33, 90.78] | 81.32 [57.33, 90.78] | 81.32 [57.58, 90.78] | 81.32 [57.44, 90.78] |
| Mean ± SD* | 96.48 ± 7.03 | 66.78 ± 41.69 | 66.78 ± 41.69 | 67.03 ± 41.21 | 66.9 ± 41.47 |

**[*]** Statistics with termination reasons held out.

For each CPET feature—peak V̇O₂, VE/VCO₂ slope, and weight—performance metrics (coverage, correct, recall, precision, and F₁) are reported separately for the regex model, few-shot prompting (FSP) model, and hybrid model. Values are presented as median [interquartile range] and mean ± standard deviation. FSP models means Few-shot prompt model.

#### Supplementary Table 9 Hybrid Model Performance Metrics

| Hybrid Model | Coverage (%) | Correct (%) | Recall (%) | Precision (%) | F1 (%) |
| --- | --- | --- | --- | --- | --- |
| peakVO₂ | 96.79 [94.6, 98.98] | 95.58 [93.03, 98.13] | 95.58 [93.03, 98.13] | 98.76 [97.36, 100.0] | 97.14 [95.06, 98.12] |
| VE/VCO2slope | 90.39 [86.58, 94.21] | 86.9 [82.53, 91.27] | 86.9 [82.53, 91.27] | 96.14 [93.51, 98.76] | 91.28 [88.05, 93.41] |
| Weight | 100.0 [100.0, 100.0] | 98.47 [96.98, 99.96] | 98.47 [96.98, 99.96] | 98.47 [96.98, 99.96] | 98.47 [96.76, 99.14] |
| TEST TYPE | 100.0 [100.0, 100.0] | 98.09 [96.43, 99.75] | 98.09 [96.43, 99.75] | 98.09 [96.43, 99.75] | 98.09 [96.32, 98.78] |
| Termination reason | 97.8 [95.4 – 99.2] | 89.3 [85.0 – 93.6] | 90.1 [87.0 – 93.2] | 94.8 [92.1 – 97.5] | 92.2 [89.6 – 96.3] |
| Median [IQR]* | 100.0 [100.0, 100.0] | 98.09 [96.43, 99.75] | 98.09 [96.43, 99.75] | 98.09 [96.43, 99.75] | 98.09 [96.44, 98.78] |
| Mean ± SD* | 98.4 [95.19 - 100.0] | 96.84 [93.41, 98.18] | 96.84 [93.41 - 98.18] | 98.28 [97.6 - 98.54] | 97.62 [95.68 - 98.18] |

**[*]** Statistics with termination reasons held out.

For each CPET feature—peak V̇O₂, VE/VCO₂ slope, and weight—performance metrics (coverage, correct, recall, precision, and F₁) are reported separately for the regex model, few-shot prompting (FSP) model, and hybrid model. Values are presented as median [interquartile range] and mean ± standard deviation. FSP models means Few-shot prompt model.

#### Supplementary Table 10 Summary of tables: Aggregated Median [IQR] and Mean ± SD of Performance Metrics for CPET Models

|  | Regex Model | | FSP Model | | hybrid model | |
| --- | --- | --- | --- | --- | --- | --- |
|  | Median  [IQR1-IQR3] | Mean  ± SD | Median  [IQR1-IQR3] | Mean  ± SD | Median  [IQR1-IQR3] | Mean  ± SD |
| Coverage (%) | 100.0  [100.00 – 100.0] | 97.19  ± 6.29 | 99.8  [97.41 – 100.0] | 97.61  ± 4.52 | 98.4  [95.19 – 100.0] | 96.8  ± 4.53 |
| Correct (%) | 74.43  [47.33 – 88.21] | 62.89  ± 37.14 | 84.44  [69.75 – 97.9] | 83.2  ± 17.35 | 96.84  [93.41 – 98.18] | 94.76  ± 5.39 |
| Recall (%) | 74.43  [50.0 – 88.21] | 63.43  ± 36.88 | 84.44  [69.75 – 97.9] | 83.2  ± 17.35 | 96.84  [93.41 – 98.18] | 94.76  ± 5.39 |
| Precision (%) | 74.43  [11.68 – 88.21] | 55.94  ± 43.35 | 88.04  [75.21 – 97.9] | 83.2  ± 17.35 | 98.28  [97.6 – 98.54] | 97.86  ± 1.18 |
| F1 (%) | 74.43  [18.00 – 88.21] | 57.12  ± 42.04 | 86.16  [72.35 – 97.9] | 84.09  ± 16.58 | 97.62  [95.68 – 98.18] | 96.24  ± 3.36 |

Coverage, Correct, Recall, Precision, and F₁ scores are reported as median [interquartile range (IQR₁–IQR₃)] and mean ± standard deviation (SD) for each model configuration: RegEx, few-shot, and hybrid. Abbreviation: CMR = cardiac magnetic resonance, FSP = Few-shot prompt model.

### Performance Comparisons for CMR and CPET Models

#### Supplementary Table 11: Performance Comparisons for CMR and CPET Models

| ComparisonS | Wilcoxon W | p-value  (Wilcoxon) | Adjusted p  p-value (Q) | Cohen's d | Cliff's delta |
| --- | --- | --- | --- | --- | --- |
| CMR Models |  |  |  |  |  |
| RegEx vs Few-shot | 36.0 | 0.55 | 0.55 | 0.03 | -0.11 |
| RegEx vs Hybrid | 0.0 | **<0.01** | 0.03 | 0.59 | 0.67 |
| Few-shot vs Hybrid | 15.0 | **0.03** | 0.045 | 0.68 | 0.67 |
| CPET Models |  |  |  |  |  |
| RegEx vs Few-shot | 1.0 | 0.14 | 0.14 | 0.76 | 0.40 |
| RegEx vs Hybrid | 0.0 | 0.06 | 0.14 | 0.91 | 0.64 |
| Few-shot vs Hybrid | 0.0 | 0.10 | 0.14 | 0.68 | 0.28 |

Note. For each model type, pairwise differences were evaluated using Wilcoxon signed‑rank tests, and p values were adjusted for multiple comparisons using the Benjamini-Hochberg false discovery rate procedure. Effect sizes are presented as Cohen’s *d* and Cliff’s δ. * indicates an FDR-adjusted *p value (q)* < .05. Abbreviations: CMR = cardiac magnetic resonance; CPET = cardiopulmonary exercise testing.

### Prompts

#### CMR Zero Shot Prompt

\### ROLE ###

You are an expert cardiac radiologist specializing in Cardiac Magnetic Resonance (CMR) report analysis.

\### TASK ###

Parse the English CMR report below and return **one single‑line JSON object** that contains **exactly** the 13 keys listed under “ALLOWED KEYS” in the specified order.

• Do **not** add extra keys, remove keys, nest objects, or output arrays/comments.

• If a value is absent in the report, set it to **null** (without quotes).

\### ALLOWED KEYS (fixed order) ###

["height","weight","BSA","LVEDV","LVESV","LVSV","LVEF","RVEDV","RVESV","RVSV","RVEF","LV Mass","PRF"]

\### EXTRACTION RULES ###

1. **Verbatim numbers only** – copy numeric values exactly as written; never derive or calculate.

2. **Strip units** – remove “%”, “cm”, “kg”, “mL/beat”, “L/min”, “m²”, “g”, etc.; keep only the number.

3. **Context by header / nearest phrase**

• Inside “LEFT VENTRICLE” → LV‑… keys • Inside “RIGHT VENTRICLE” → RV‑… keys • Aortic “regurgitant fraction” → ARF • Pulmonary/RVOT “regurgitant fraction” → PRF

4. **Synonyms to keys**

HT/height, WT/weight, BSA. EDV/ESV/SV/EF map to LV‑ or RV‑ keys per context.

“LV mass” maps to “LV Mass” (non‑indexed) or “LVMi” when indexed.

5. **Ignore** qualitative words (“mild”, “severe”), measurements not in the 13 keys, narrative descriptions, imaging parameters, anatomical findings outside scope.

6. **Ambiguity** – if you cannot unambiguously assign LV vs. RV or any key, output **null** for that key.

7. **Output format** – one single‑line JSON starting with “{” and ending with “}”, keys strictly in the order shown above.

\### REPORT ###

$report

\### REQUIRED OUTPUT ###

A single‑line JSON object with the 13 keys in the order given.

#### CMR One Shot Prompt

\### ROLE ###

You are an expert cardiac radiologist specializing in Cardiac Magnetic Resonance (CMR) report analysis.

\### TASK ###

Parse an English CMR report and output a single JSON object with exactly the 13 keys listed below, in the specified order. Do not include any additional keys or omit any listed keys. If a value is not explicitly stated in the report, set it to null. The output must be a single-line JSON object starting with { and ending with }.

\### ALLOWED KEYS (fixed order) ###

["height","weight","BSA","LVEDV","LVESV","LVSV","LVEF","RVEDV","RVESV","RVSV","RVEF","LV Mass","PRF"]

\### EXTRACTION RULES ###

1. **Verbatim numbers only** – copy numeric values exactly as written; never derive or calculate.

2. **Strip units** – remove “%”, “cm”, “kg”, “mL/beat”, “L/min”, “m²”, “g”, etc.; keep only the number.

3. **Context by header / nearest phrase**

• Inside “LEFT VENTRICLE” → LV‑… keys • Inside “RIGHT VENTRICLE” → RV‑… keys • Aortic “regurgitant fraction” → ARF • Pulmonary/RVOT “regurgitant fraction” → PRF

4. **Synonyms to keys**

HT/height, WT/weight, BSA. EDV/ESV/SV/EF map to LV‑ or RV‑ keys per context.

“LV mass” maps to “LV Mass” (non‑indexed) or “LVMi” when indexed.

5. **Ignore** qualitative words (“mild”, “severe”), measurements not in the 13 keys, narrative descriptions, imaging parameters, anatomical findings outside scope.

6. **Ambiguity** – if you cannot unambiguously assign LV vs. RV or any key, output **null** for that key.

7. **Output format** – one single‑line JSON starting with “{” and ending with “}”, keys strictly in the order shown above.

EXAMPLE INPUT AND OUTPUT

Input Report 1:

OUTFLOW: There is no right ventricular outflow tract obstruction (peak velocity = 1.3 m/s). There is an intact pulmonary valve. There is severe pulmonary regurgitation (regurgitant fraction = 47%, regurgitant volume = 38 mL/beat or 25 mL/beat/m2).

There is no left ventricular outflow tract obstruction. The aortic valve is trileaflet with thickening of each leaflet, with trivial stenosis (peak velocity = 1.8 m/s) and mild regurgitation (regurgitant fraction = 13%).

HT: 158 cm WT: 50.9 kg BSA: 1.5 m2

RIGHT VENTRICLE:

Ejection Fraction = 46%, End Diastolic Volume = 119 mL/m2, End Systolic Volume = 64 ml/m2, Stroke Volume = 83 mL, Myocardial Mass = 21 g/m2.

LEFT VENTRICLE:

Ejection Fraction = 51%, End Diastolic Volume = 103 mL/m2, End Systolic Volume = 51 ml/m2, Stroke Volume = 78 mL, Myocardial Mass = 38 g/m2.

Output:

{"height":158,"weight":50.9,"BSA":1.5,"LVEDV":null,"LVESV":null,"LVSV":78,"LVEF":51,"RVEDV":null,"RVESV":null,"RVSV":83,"RVEF":46, "LV Mass":null, "PRF":47}

\### NOW PROCESS THE FOLLOWING REPORT ###

$report

Output exactly one JSON object with the 13 keys in the specified order.

#### CMR Few Shot Prompt

\### ROLE ###

You are an expert cardiac radiologist specializing in Cardiac Magnetic Resonance (CMR) report analysis.

\### TASK ###

Parse an English CMR report and output a single JSON object with exactly the 13 keys listed below, in the specified order. Do not include any additional keys or omit any listed keys. If a value is not explicitly stated in the report, set it to null. The output must be a single-line JSON object starting with { and ending with }.

\### ALLOWED KEYS (fixed order) ###

["height","weight","BSA","LVEDV","LVESV","LVSV","LVEF","RVEDV","RVESV","RVSV","RVEF","LV Mass","PRF"]

\### EXTRACTION RULES ###

1 Extract only explicit numerical values: Copy numbers as they appear in the report. Do not calculate, infer, or derive values (e.g., do not compute stroke volume from EDV and ESV).

2 Strip units: Remove units such as “%”, “cm”, “kg”, “mL”, “cc”, “g”, “m²”, “L/min”, “mL/beat”, etc., and output only the numerical value.

3  Context → use nearest header/phrase:

• Values in “LEFT VENTRICLE” block → LV‑… keys.

• Values in “RIGHT VENTRICLE” block → RV‑… keys.

• “regurgitant fraction” under pulmonary valve/artery/RVOT → PRF.

4  Synonyms: HT/height, WT/weight, BSA. EDV/ESV/SV/EF map to LV‑ or RV‑ keys per context. “LV mass” → LV Mass or LVMi per “index”.

5 Context awareness: Use section headers (e.g., "LEFT VENTRICLE", "RIGHT VENTRICLE", "OUTFLOW", "FLOW QUANTIFICATION") or surrounding text to determine whether a value pertains to LV, RV, pulmonary artery. For example:

- "EF" under "LEFT VENTRICLE" maps to LVEF.

- "regurgitant fraction" under "PULMONARY VALVE" or "main pulmonary artery" maps to PRF.

6 Ignore irrelevant data: Disregard:

- Qualitative descriptions (e.g., "mild", "severe", "dilated").

- Measurements not listed in the 13 keys (e.g., regurgitant volume, peak velocity, pulmonary artery dimensions, aortic root diameter).

- Narrative text, section headers, metadata, tables, and imaging details (e.g., "mDixon sequence", "Siemens Avanto").

- Other anatomical details (e.g., coronary artery origins, pleural thickening, VSD patch).

7 Handle ambiguity: If a value’s context (e.g., LV vs. RV) is unclear or the measurement is not explicitly tied to one of the 13 keys, set it to null. Do not guess.

8 Output format: Ensure the output is a single-line JSON object with exactly the 13 keys. Do not include nested objects, arrays, comments, or extra fields.

EXAMPLE INPUT AND OUTPUT

Input Report 1:

OUTFLOW: There is no right ventricular outflow tract obstruction (peak velocity = 1.3 m/s). There is an intact pulmonary valve. There is severe pulmonary regurgitation (regurgitant fraction = 47%, regurgitant volume = 38 mL/beat or 25 mL/beat/m2).

There is no left ventricular outflow tract obstruction. The aortic valve is trileaflet with thickening of each leaflet, with trivial stenosis (peak velocity = 1.8 m/s) and mild regurgitation (regurgitant fraction = 13%).

HT: 158 cm WT: 50.9 kg BSA: 1.5 m2

RIGHT VENTRICLE:

Ejection Fraction = 46%, End Diastolic Volume = 119 mL/m2, End Systolic Volume = 64 ml/m2, Stroke Volume = 83 mL, Myocardial Mass = 21 g/m2.

LEFT VENTRICLE:

Ejection Fraction = 51%, End Diastolic Volume = 103 mL/m2, End Systolic Volume = 51 ml/m2, Stroke Volume = 78 mL, Myocardial Mass = 38 g/m2.

Output:

{"height":158,"weight":50.9,"BSA":1.5,"LVEDV":null,"LVESV":null,"LVSV":78,"LVEF":51,"RVEDV":null,"RVESV":null,"RVSV":83,"RVEF":46, "LV Mass":null,"PRF":47}

Input Report 2:

LEFT VENTRICLE: Normal size, wall thickness and low normal systolic function. LV volumetric parameters as follows: EF: 50 % EDV: 132 ml ESV: 66 ml SV: 66 ml CO: 5.7 L/min

RIGHT VENTRICLE: Dilated with mild to moderate systolic dysfunction. RV volumetric parameters as follows: EF: 32 % EDV: 174 ml ESV: 119 ml SV: 55 ml CO: 4.8 L/min

Flow quantification through the ascending aorta: Aortic regurgitant fraction = 18 %

Output:

{"height":null,"weight":null,"BSA":null,"LVEDV":132,"LVESV":66,"LVSV":66,"LVEF":50,"RVEDV":174,"RVESV":119,"RVSV":55,"RVEF":32, "LV Mass":null,"PRF":null}

Input Report 3:

Left ventricle: EDV = 155 cc; EDVi = 78 cc/m2. ESV = 63 cc; ESVi = 32 cc/m2. Stroke volume = 92 cc; SVi = 46 cc/m2. LVEF = 60 %. Cardiac Output = 6.0 l/min.; Cardiac Index = 3.0 l/min/m2. LV mass = 69 gm; LVMi = 35 gm/m2.

Right ventricle: EDV = 195 cc; EDVi = 98 cc/m2. ESV = 117 cc; ESVi = 59 cc/m2. Stroke volume = 78 cc; SVi = 39 cc/m2. RVEF = 40 %.

Pulmonic regurgitant fraction: 11%

Qp/Qs = 1.04

Output:

{"height":null,"weight":null,"BSA":null,"LVEDV":155,"LVESV":63,"LVSV":92,"LVEF":60,"RVEDV":195,"RVESV":117,"RVSV":78,"RVEF":40 ,"LV Mass":69,"PRF":11}

\### NOW PROCESS THE FOLLOWING REPORT ###

$report

Output exactly one JSON object with the 13 keys in the specified order.

#### CPET Zero Shot Prompt

\### ROLE ###

You are an expert in analyzing Cardiopulmonary Exercise Testing (CPET) reports. Your task is to extract specific numerical and categorical data from these reports based on the provided instructions.

---

\### TASK ###

Parse the given CPET report and extract the following six pieces of information in the specified order. If a value is not explicitly stated in the report, set it to `null`. The output must be a single-line JSON object starting with `{` and ending with `}`.

---

\### REQUIRED KEYS (in fixed order) ###

1. `weight_kg` – Patient's weight in kilograms.

2. `peak_vo2` – Peak VO2 value (in ml/kg/min).

3. `ve_vco2_slope` – VE/VCO2 slope value.

4. `test_mode` – The mode of the test (e.g., "treadmill" or "cycle").

5. `absolute_termination_reason` – The reason for test termination that matches **absolute** criteria; otherwise, `null`.

6. `relative_termination_reason` – The reason for test termination that matches **relative** criteria; otherwise, `null`.

---

\### EXTRACTION RULES ###

1. **Explicit Values Only**: Extract only explicitly stated numerical or categorical values. Do not infer or calculate values.

2. **Units**: Remove units (e.g., "ml/kg/min", "kg") and output only the numerical value.

3. **Context Awareness**:

- `weight_kg`: Extract from the demographic section (e.g., “WEIGHT: 72.3 kgs”).

- `peak_vo2`: From the **IMPRESSION** section, only if “Peak” appears on the same line; prefer ml/kg/min; ignore everything after “Treadmill Summary.”

- `ve_vco2_slope`: Only if explicitly mentioned as “VE/VCO2 slope: X” or “slope (X, …)”.

- `test_mode`: Extract from “TEST MODE” field.

- `absolute_termination_reason`: Only absolute stop criteria (e.g., angina, syncope, VT, ST elevation).

- `relative_termination_reason`: Only relative stop criteria (e.g., fatigue, dyspnea, minor ST changes).

4. **Handle Ambiguity**: If unclear or missing, use `null`.

5. **Output Format**: Single-line JSON with the six keys in the specified order.

---

\### NOW PROCESS THE FOLLOWING REPORT ###

The full report is below:

$narrative_full

Output exactly one JSON object with the five keys in the specified order.

#### CPET One Shot Prompt

\### ROLE ###

You are an expert in analyzing Cardiopulmonary Exercise Testing (CPET) reports. Your task is to extract specific numerical and categorical data from these reports based on the provided instructions.

---

\### TASK ###

Parse the given CPET report and extract the following six pieces of information in the specified order. If a value is not explicitly stated in the report, set it to `null`. The output must be a single-line JSON object starting with `{` and ending with `}`.

---

\### REQUIRED KEYS (in fixed order) ###

1. `weight_kg` – Patient's weight in kilograms.

2. `peak_vo2` – Peak VO2 value (in ml/kg/min).

3. `ve_vco2_slope` – VE/VCO2 slope value.

4. `test_mode` – The mode of the test (e.g., "treadmill" or "cycle").

5. `absolute_termination_reason` – The reason for test termination that matches **absolute** criteria (if any); otherwise, `null`.

6. `relative_termination_reason` – The reason for test termination that matches **relative** criteria (if any); otherwise, `null`.

---

\### EXTRACTION RULES ###

1. **Explicit Values Only**: Extract only explicitly stated numerical or categorical values. Do not infer or calculate values.

2. **Units**: Remove units (e.g., "ml/kg/min", "kg", "cm", "L/min") and output only the numerical value.

3. **Context Awareness**:

    - `weight_kg`: Extract from the demographic section (e.g., "WEIGHT: 72.3 kgs").

    - `peak_vo2`: This refers to the narratively reported peak oxygen consumption, that must be found in the **IMPRESSION** section. Only extract the value if the word “Peak” appears in the same line (e.g., “Peak VO2 (1.91 l/min) and abnormal low VO2/kg 24.3 ml/min/kg”). Prefer values reported in ml/kg/min. Ignore everything after **Treadmill Summary**, even if they show a “VO2Max” or “ml/kg/min” number.

    - `ve_vco2_slope`: Extract only if the slope is explicitly mentioned in the narrative text, such as:

    “VE/VCO2 slope: 26”

    “VE/VCO2 at AT and slope (32, Z=-0.03)”

     Do not use VE/VCO2 values from individual test stages (AT or VO2Max) in tables.

   - `test_mode`: Extract from "TEST MODE" or similar fields.

   - `absolute_termination_reason`: Extract from statements indicating test stopped due to **absolute** criteria such as:

     - Significant drop in SBP with ischemia

     - Moderate-to-severe angina

     - Neurologic symptoms like dizziness, ataxia, near-syncope

     - Cyanosis or pallor

     - Ventricular tachycardia

     - ST elevation

   - `relative_termination_reason`: Extract from statements indicating **relative** stop criteria such as:

     - Fatigue, lightheadedness, shortness of breath

     - Supraventricular arrhythmias, PVCs, bradycardia

     - Excessive BP (SBP ≥ 250 or DBP ≥ 115)

     - Minor ST or QRS changes

   If both types apply, list them in the correct fields. If no clear cause is given, set both to `null`.

4. **Handle Ambiguity**: If the value or classification is unclear or ambiguous, set to `null`.

5. **Output Format**: Return a single-line JSON object with the six keys in the **specified order**.

---

\### EXAMPLE INPUT AND OUTPUT ###

\#### Example Input Report 1:

Height: 180.50 Cms  Weight: 72.3 Kgs

... TEST MODE: TREADMILL

IMPRESSION:

... Metabolic Exercise Test Report: abnormal or normal PEAK VO2: (2.45 l/min) and normal or abnormal low VO2/kg 18.1 ml/kg/min

Normal VE/VCO2 at AT and slope (29, Z=-0.03)

Treadmill Summary:

Middle 5 of 7 Mean

                                  Rest     AT    VO2Max  PredMax AT/Max  Max/Pred

WORK

VO2 (mL/kg/min)                     6.5    17.5    27.1    47.6      37      57

... OBSERVATION:

The test was terminated due to dizziness and shortness of breath.

\#### Example Output:

{"weight_kg":72.3,"peak_vo2":18.1,"ve_vco2_slope":29,"test_mode":"treadmill","absolute_termination_reason":null,"relative_termination_reason":"dizziness and shortness of breath"}

\### NOW PROCESS THE FOLLOWING REPORT ###

The full report is below:

$narrative_full

Output exactly one JSON object with the five keys in the specified order.

#### CPET Few Shot Prompt

\### ROLE ###

You are an expert in analyzing Cardiopulmonary Exercise Testing (CPET) reports. Your task is to extract specific numerical and categorical data from these reports based on the provided instructions.

---

\### TASK ###

Parse the given CPET report and extract the following six pieces of information in the specified order. If a value is not explicitly stated in the report, set it to `null`. The output must be a single-line JSON object starting with `{` and ending with `}`.

---

\### REQUIRED KEYS (in fixed order) ###

1. `weight_kg` – Patient's weight in kilograms.

2. `peak_vo2` – Peak VO2 value (in ml/kg/min).

3. `ve_vco2_slope` – VE/VCO2 slope value.

4. `test_mode` – The mode of the test (e.g., "treadmill" or "cycle").

5. `absolute_termination_reason` – The reason for test termination that matches **absolute** criteria (if any); otherwise, `null`.

6. `relative_termination_reason` – The reason for test termination that matches **relative** criteria (if any); otherwise, `null`.

---

\### EXTRACTION RULES ###

1. **Explicit Values Only**: Extract only explicitly stated numerical or categorical values. Do not infer or calculate values.

2. **Units**: Remove units (e.g., "ml/kg/min", "kg", "cm", "L/min") and output only the numerical value.

3. **Context Awareness**:

    - `weight_kg`: Extract from the demographic section (e.g., "WEIGHT: 72.3 kgs").

    - `peak_vo2`: This refers to the narratively reported peak oxygen consumption, that must be found in the **IMPRESSION** section. Only extract the value if the word “Peak” appears in the same line (e.g., “Peak VO2 (1.91 l/min) and abnormal low VO2/kg 24.3 ml/min/kg”). Prefer values reported in ml/kg/min. Ignore everything after **Treadmill Summary**, even if they show a “VO2Max” or “ml/kg/min” number.

    - `ve_vco2_slope`: Extract only if the slope is explicitly mentioned in the narrative text, such as:

    “VE/VCO2 slope: 26”

    “VE/VCO2 at AT and slope (32, Z=-0.03)”

     Do not use VE/VCO2 values from individual test stages (AT or VO2Max) in tables.

   - `test_mode`: Extract from "TEST MODE" or similar fields.

   - `absolute_termination_reason`: Extract from statements indicating test stopped due to **absolute** criteria such as:

     - Significant drop in SBP with ischemia

     - Moderate-to-severe angina

     - Neurologic symptoms like dizziness, ataxia, near-syncope

     - Cyanosis or pallor

     - Ventricular tachycardia

     - ST elevation

   - `relative_termination_reason`: Extract from statements indicating **relative** stop criteria such as:

     - Fatigue, lightheadedness, shortness of breath

     - Supraventricular arrhythmias, PVCs, bradycardia

     - Excessive BP (SBP ≥ 250 or DBP ≥ 115)

     - Minor ST or QRS changes

   If both types apply, list them in the correct fields. If no clear cause is given, set both to `null`.

4. **Handle Ambiguity**: If the value or classification is unclear or ambiguous, set to `null`.

5. **Output Format**: Return a single-line JSON object with the six keys in the **specified order**.

---

\### EXAMPLE INPUT AND OUTPUT ###

\#### Example Input Report 1:

Height: 180.50 Cms  Weight: 72.3 Kgs

... TEST MODE: TREADMILL

IMPRESSION:

... Metabolic Exercise Test Report: abnormal or normal PEAK VO2: (2.45 l/min) and normal or abnormal low VO2/kg 18.1 ml/kg/min

Normal VE/VCO2 at AT and slope (29, Z=-0.03)

Treadmill Summary:

Middle 5 of 7 Mean

                                  Rest     AT    VO2Max  PredMax AT/Max  Max/Pred

WORK

VO2 (mL/kg/min)                     6.5    17.5    27.1    47.6      37      57

... OBSERVATION:

The test was terminated due to dizziness and shortness of breath.

\#### Example Output:

{"weight_kg":72.3,"peak_vo2":18.1,"ve_vco2_slope":29,"test_mode":"treadmill","absolute_termination_reason":null,"relative_termination_reason":"dizziness and shortness of breath"}

\#### Example Input Report 2:

WEIGHT: 72.3 kg/159.1 lbs, HEIGHT: 157.5 cm/62.0 inches

METABOLIC EXERCISE TEST REPORT

DATA SUMMARY

Peak RER: 1.17

Peak VO2: 63 % of predicted (18.1 ml/kg/min)

...

VE/VCO2 slope: 29

\#### Example Output:

{"weight_kg":72.3,"peak_vo2":18.1,"ve_vco2_slope":29,"test_mode":null,"absolute_termination_reason":null,"relative_termination_reason":null}

\#### Example Input Report 3:

Metabolic Exercise Test Report:

Peak VO2 (30 sec avg) = 26.0 ml/kg/min at R = 1.13 (adequate effort).

Ventilatory anaerobic threshold …

Rapid Respiratory Rate during exercise …

(Oxygen-saturation lines, NYHA class, etc.)

{"weight_kg":null,"peak_vo2":26.0,"ve_vco2_slope":null,"test_mode":null,"absolute_termination_reason":null,"relative_termination_reason":null}

\### NOW PROCESS THE FOLLOWING REPORT ###

The full report is below:

$narrative_full

Output exactly one JSON object with the five keys in the specified order.

### Regex Patterns

**Heights:** "(?i)\b(?:height|ht)\b\s*[:=]\s*(\d+(?:\.\d+)?)\s*(?:cm|centimeters)?"

**Weights:** "(?i)\b(?:weight|wt)\b\s*[:=]\s*(\d+(?:\.\d+)?)\s*(?:kg|kilograms)?"

**BSA:** "(?i)\b(?:bsa|body\s+surface\s+area)\b\s*[:=]\s*(\d+(?:\.\d+)?)\s*(?:m2|m²)?"

**LV:** "(?is)left\s+ventricle[\s\S]*?(?=right\s+ventricle|$)"

**RV:** "(?is)right\s+ventricle[\s\S]*?(?=left\s+ventricle|$)"

**EDV:** "(?i)\b(?:EDV|End\s+Diastolic\s+Volume)\b[^:=]*[:=]\s*(\d+(?:\.\d+)?)"

**ESV:** "(?i)\b(?:ESV|End\s+Systolic\s+Volume)\b[^:=]*[:=]\s*(\d+(?:\.\d+)?)"

**SV:** "(?i)\b(?:SV|Stroke\s+Volume)\b[^:=]*[:=]\s*(\d+(?:\.\d+)?)"

**EF:** "(?i)\b(?:lv\s*ef|rv\s*ef|[lr]?vef|ejection\s+fraction)(?:\s*\(total\))?[^:=]*[:=]\s*(\d+(?:\.\d+)?)(?:\s*\%?)"

**LVMass:** "(?i)\b(?:LV\s*mass|Myocardial\s+Mass)\b[^:=]*[:=]\s*(\d+(?:\.\d+)?)"

**RVMass:** "(?i)\b(?:RV\s*mass|Myocardial\s+Mass)\b[^:=]*[:=]\s*(\d+(?:\.\d+)?)"

**PRF:** "(?i)\b(?:pulmonary\s+regurgitant\s+fraction|pulmonary\s+regurgitation\s+fraction|prf)\b[^:=]*[:=]\s*(\d+(?:\.\d+)?)(?:\s*\%?)"

**peakVO2:** “\bpeak\W+vo2[^0-9]{0,80}?(?:=|:|\()?\s*(?P<peak_vo2>\d+(?:\.\d+)?) \s*(?:ml\s*/\s*kg\s*/\s*min|l\s*/\s*min)”

**VE/VCO2:** “\bve\s*\/\s*vco2(?:\s*(?:slope)?)?[^0-9]{0,80}?(?P<ve_vco2>\d+(?:\.\d+)?)”

**TEST TERMINATION:** "(?P<termination>(?:\btest\s+(?:was\s+)?terminated\b|\btest\s+(?:was\s+)?stopped\b|\bterminated\s+due\s+to\b|\bterminated\s+because\b|\bstop(?:ped)?\s+due\s+to\b|\bend\s+criteria\b|\breason\s+for\s+test\s+termination\b|\bbreakreason\s*\d+\s*[A-Za-z]\b)[^.\n]{0,120})"
